# A novel cortical biomarker signature predicts individual pain sensitivity

**DOI:** 10.1101/2024.06.16.24309005

**Authors:** Nahian S Chowdhury, Chuan Bi, Andrew J Furman, Alan KI Chiang, Patrick Skippen, Emily Si, Samantha K Millard, Sarah M Margerison, Darrah Spies, Michael L Keaser, Joyce T Da Silva, Shuo Chen, Siobhan M Schabrun, David A Seminowicz

**Affiliations:** Center for Pain IMPACT, Neuroscience Research Australia, Sydney, New South Wales, Australia; University of New South Wales, Sydney, New South Wales, Australia; Division of Biostatistics and Bioinformatics, Department of Epidemiology and Public Health, University of Maryland School of Medicine; Division of Biostatistics, Center for Devices and Radiological Health, U.S. Food and Drug Administration, USA; Department of Neural and Pain Sciences, University of Maryland School of Dentistry, Baltimore, USA; Center to Advance Chronic Pain Research, University of Maryland Baltimore, USA; Data Sciences, Hunter Medical Research Institute, Newcastle, NSW, Australia; School of Medicine and Public Health, University of Newcastle, Newcastle, NSW, Australia; The Gray Centre for Mobility and Activity, Parkwood Institute, St. Joseph’s Healthcare London, Canada; School of Physical Therapy, University of Western Ontario, London, Canada; Department of Medical Biophysics, Schulich School of Medicine & Dentistry, University of Western Ontario, London, Canada

**Author notes:** Co-first author (Nahian S Chowdhury, Chuan Bi). Co-senior author (Siobhan M Schabrun, David A Seminowicz). **Corresponding Author:** Professor David A Seminowicz Department of Medical Biophysics Schulich School of Medicine & Dentistry University of Western Ontario, London, Canada.

**Keywords:** Pain, Biomarker, Electroencephalography, Transcranial Magnetic Stimulation

## Abstract

**Importance:** Biomarkers would greatly assist decision making in the diagnosis, prevention and treatment of chronic pain.

**Objective:** The present study aimed to undertake analytical validation of a sensorimotor cortical biomarker signature for pain consisting of two measures: sensorimotor peak alpha frequency (PAF) and corticomotor excitability (CME).

**Design:** In this cohort study (recruitment period: November 2020-October 2022), participants experienced a model of prolonged temporomandibular pain with outcomes collected over 30 days. Electroencephalography (EEG) to assess PAF and transcranial magnetic stimulation (TMS) to assess CME were recorded on Days 0, 2 and 5. Pain was assessed twice daily from Days 1-30.

**Setting:** Data collection occurred at a single centre: Neuroscience Research Australia.

**Participants:** We enrolled 159 healthy participants (through notices placed online and at universities across Australia), aged 18-44 with no history of chronic pain, neurological or psychiatric condition. 150 participants completed the protocol.

**Exposure:** Participants received an injection of nerve growth factor (NGF) to the right masseter muscle on Days 0 and 2 to induce prolonged temporomandibular pain lasting up to 4 weeks.

**Main Outcomes and Measures:** We determined the predictive accuracy of the PAF/CME biomarker signature using a nested control-test scheme: machine learning models were run on a training set (n = 100), where PAF and CME were predictors and pain sensitivity was the outcome. The winning classifier was assessed on a test set (n = 50) comparing the predicted pain labels against the true labels.

**Results:** The final sample consisted of 66 females and 84 males with a mean age of 25.1 ± 6.2. The winning classifier was logistic regression, with an outstanding area under the curve (AUC=1.00). The locked model assessed on the test set had excellent performance (AUC=0.88[0.78-0.99]). Results were reproduced across a range of methodological parameters. Moreover, inclusion of sex and pain catastrophizing as covariates did not improve model performance, suggesting the model including biomarkers only was more robust. PAF and CME biomarkers showed good-excellent test-retest reliability.

**Conclusions and Relevance:** This study provides evidence for a sensorimotor cortical biomarker signature for pain sensitivity. The combination of accuracy, reproducibility, and reliability, suggests the PAF/CME biomarker signature has substantial potential for clinical translation, including predicting the transition from acute to chronic pain.

**Key Points:** *Question:* Can individuals be accurately classified as high or low pain sensitive based on two features of cortical activity: sensorimotor peak alpha frequency (PAF) and corticomotor excitability (CME)?

*Findings:* In a cohort study of 150 healthy participants, the performance of a logistic regression model was outstanding in a training set (n=100) and excellent in a test set (n=50), with the combination of slower PAF and CME depression predicting higher pain. Results were reproduced across a range of methodological parameters.

*Meaning:* A novel cortical biomarker can accurately distinguish high and low pain sensitive individuals, and may predict the transition from acute to chronic pain

---

Several objective pain biomarkers have been proposed, including neuroimaging markers of mechanistic/structural abnormalities^1-3^, neural oscillatory rhythms^4^ and “multi-omics” metrics of micro RNA^5^, proteins^6^, lipids and metabolites^7^. Such biomarkers would greatly assist decision making in the diagnosis, prevention and treatment of chronic pain^8^. However, attempts at establishing pain biomarkers have suffered from either insufficient sample sizes to conduct full-scale analytical validation using machine learning^8-10^, failure to use clinically relevant pain models^11-13^ or lack of assessment of reproducibility or test-retest reliability^14,15^. These factors have hindered the clinical translatability of prospective pain biomarkers.

Research suggests that the neural oscillatory rhythms involved in processing nociceptive input, and the corticospinal signalling involved in the subsequent motor response, are both critical in shaping the subjective experience of pain^4,16^. This work has culminated to the identification of a promising sensorimotor cortical biomarker signature for predicting pain sensitivity involving two metrics: 1) sensorimotor peak alpha frequency (PAF), defined as the dominant sensorimotor cortical oscillation in the alpha (8-12Hz) range^17^ and 2) corticomotor excitability (CME), defined as the efficacy at which signals are relayed from primary motor cortex (M1) to peripheral muscles^18^. Previous work has shown that slower PAF prior to pain onset and reduced CME during prolonged pain (“depression”) are associated with more pain, while faster PAF and increased CME (“facilitation”) are associated with less pain^19-23^. Given individuals who experience higher pain in the early stages of a prolonged pain episode (e.g. post-surgery) are more likely to develop chronic pain in the future^24^, slow PAF prior to an anticipated prolonged pain episode and/or CME depression during the acute stages of pain could be predictors for the transition to chronic pain.

This paper presents the main outcomes of the PREDICT trial, a pre-registered (NCT04241562^25^) analytical validation of the PAF/CME biomarker signature using a model of prolonged myofascial temporomandibular pain (masseter intramuscular injection of nerve growth factor [NGF]). Repeated NGF injections induce progressively developing prolonged pain lasting up to 4 weeks^23,26^, and has been shown to mimic chronic pain characteristics such as time course (gradual development), type of pain (movement-evoked), functional impairments, hyperalgesia (decreased pressure pain thresholds) and mechanism of sensitization^27,28^. This makes the NGF model a highly standardised prolonged pain model with which to undertake biomarker validation.

The aim of the PREDICT trial was to determine whether individuals could be accurately classified as high or low pain sensitive based on baseline PAF and CME facilitator/depressor classification. We predicted the area under the curve (AUC) of the receiver operator characteristic (ROC) curve for distinguishing high and low pain sensitive individuals would be at least 70% (which represents an acceptable AUC)^29^.

## Methods

### Participants

The PREDICT trial enrolled 159 healthy participants (70 females, 89 males, mean age 25.1 ± 6.1), with 150 participants remaining after dropouts. Ethical approval was obtained from the University of New South Wales (HC190206) and the University of Maryland Baltimore (HP-00085371). Written, informed consent was obtained. The supplementary appendix contains all additional details regarding participant characteristics and methodology. We followed the “Strengthening the Reporting of Observational Studies in Epidemiology” (STROBE) reporting guidelines for cohort studies.

### Experimental Protocol

Outcomes were collected over a period of 30 days. Participants attended the laboratory on Day 0, 2, and 5. Baseline questionnaire data were collected on Day 0. Pressure pain thresholds, PAF and CME were measured on Day 0, 2 and 5. PAF was obtained via a 5-minute eyes-closed resting-state EEG recording from 63 electrodes. Sensorimotor PAF was computed by identifying the component in the signal (transformed by independent component analysis) that had a clear alpha peak (8-12Hz) upon frequency decomposition and a scalp topography suggestive of a sensorimotor source. CME was obtained using transcranial magnetic stimulation (TMS) mapping; single pulses of TMS delivered to the left primary motor cortex (M1), and motor evoked potentials (MEPs) recorded from the right masseter muscle using electromyography (EMG) electrodes. TMS was delivered at each site on a 1cm-spaced grid superimposed over the scalp, and a map of the corticomotor representation of the masseter muscle was generated. Corticomotor excitability was indexed as map volume, which is calculated by summing MEP amplitudes from all “active sites” on the grid. NGF was injected into the right masseter muscle at the end of the Day 0 and 2 laboratory sessions. Electronic pain diaries were collected from Days 1 to 30 at 10am and 7pm each day, where participant rated their pain (0-10) during various activities. Pain upon functional jaw movement is a key criterion for the diagnosis of TMD^30^ and pain during chewing and yawning are higher compared to other activities after an NGF injection to the masseter muscle^28,31^. As such, the primary outcomes were pain upon chewing and yawning. The protocol and methodology are shown in Figure 1A and 1B.

**Figure 1.**
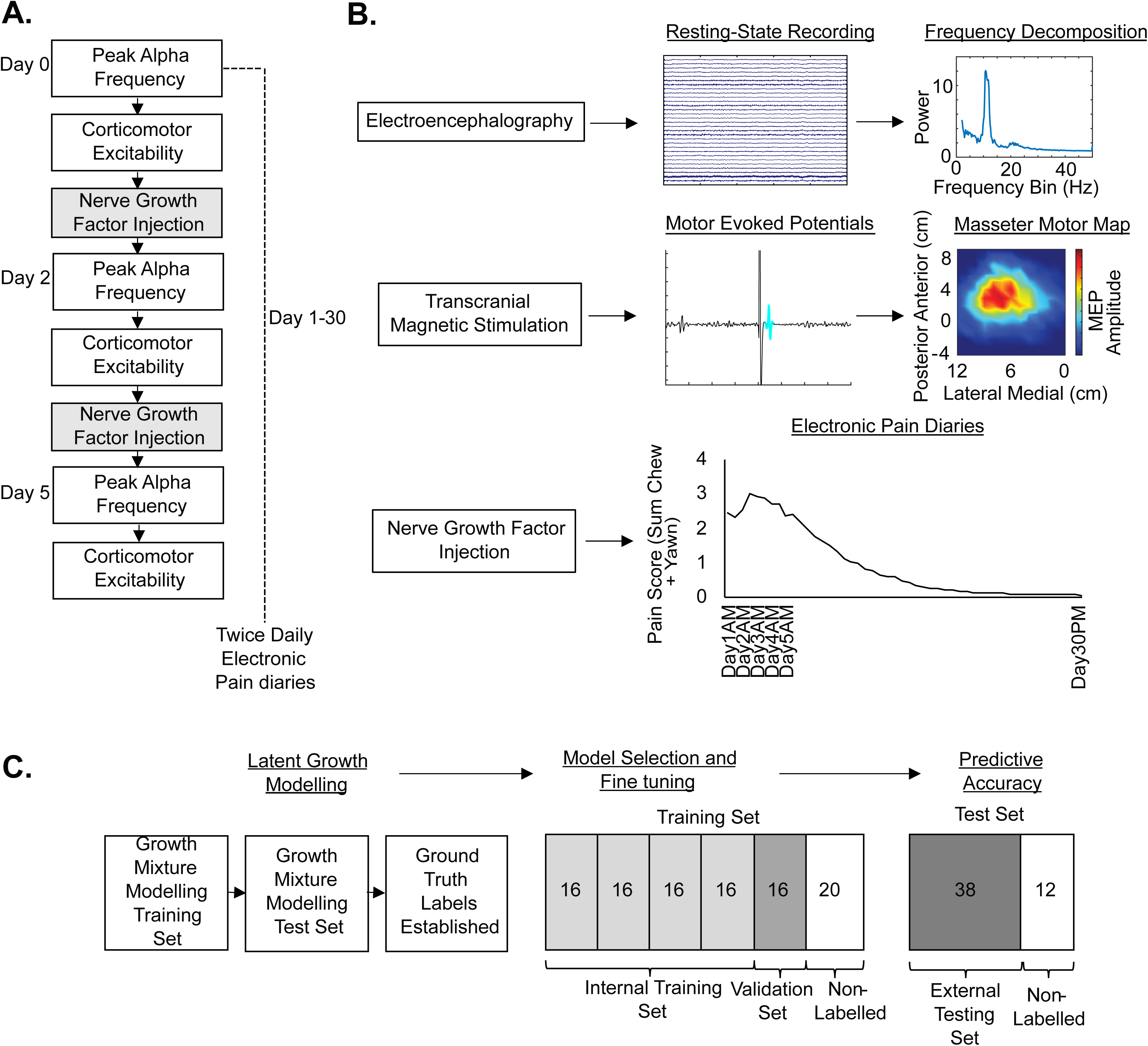
Study methodology, including study design, data collection procedures and analysis plan. **(A)** Experimental protocol showing timeline of data collection procedures. On Day 0, we measured peak alpha frequency (PAF) and corticomotor excitability (CME). At the end of the session, an injection of nerve growth factor (NGF) was administered to the right masseter muscle. On Day 2, PAF and CME were measured, followed by a second NGF injection. On Day 5, PAF and CME were measured. From Days 1-30, electronic diaries measuring jaw pain were sent to participants at 10AM and 7PM each day. **(B)** Details of the methodology. Sensorimotor PAF was measured using a 5 minutes eyes closed resting state EEG recording. Sensorimotor PAF was computed by identifying the component in the signal (transformed by independent component analysis) that had a clear alpha peak in the 8–12 Hz range upon frequency decomposition and a scalp topography suggestive of a source predominately over the sensorimotor cortex. TMS mapping was conducted by stimulating the scalp area over left M1 to obtain a map of the representation of the right masseter muscle. The map consists of the motor-evoked potential (MEP) amplitude at each stimulated location, with CME corresponding to the map volume (sum of all MEPs from active sites). Pain data was collected via electronic diaries sent to participants at 10am and 7pm each day for 30 days. The Pain score was quantified as the sum of scores for pain upon chewing and yawning. **(C)** Details of the analysis plan. We adopted a nested-control-test scheme by partitioning the 150 subjects into a training set consisting of 100 subjects and an independent test set of 50 subjects. We labelled a subset of participants in the training (n = 80) and test set (n = 38) as high or low pain sensitive using growth mixture modelling (GMM) to establish “ground-truth” labels. We then ran various machine learning models on the labelled training set (with PAF/CME as predictors, and pain severity labels as outcome), and determined optimized parameters through 5-fold cross-validation i.e. randomly dividing the 80 subjects into an internal training set of 64 subjects (with 4 equal folds of 16) and a validation set of 16. The optimized models in the internal training set were employed to predict labels in the validation set to facilitate model selection. The model with the best performance on the validation set was then locked in, and applied to the labelled test set, comparing the predicted labels of high/low pain sensitive with the ground-truth labels of high/low pain sensitive.

### Statistical Analysis

#### Division of the Data

Analysis was conducted in R, MATLAB and Python, with code publicly available https://github.com/DrNahianC/PREDICT_Scripts. Figure 1C details the analysis plan. We adopted a nested-control-testing scheme by partitioning 150 participants into the first 100 (training set) and second 50 (test set) individuals to participate in the study.

#### Growth Mixture Modelling

We used growth mixture modelling (GMM) in R^32-34^ to form two classes: high and low pain sensitive. For this categorization, we used the sum of pain upon chewing and yawning data, and pain data from Days 1-7, as this was the timeframe when pain was most prominent (eFigure 3). As such, participants would more reliably fall into high and low pain sensitive classes during this timeframe. The first and last 40 participants (80 in total) in the training set, based on the ordering of probabilities of the pain intensity trajectory belonging to one of the classes, were labelled as high and low pain sensitive. The trained GMM model, once established, was locked and utilized to label the test set. Consequently 38 out of 50 test set participants were labelled, with a different proportion of high and low pain sensitive (24 high and 14 low pain) compared to the training set since the classifications were based on the probability thresholds established in the training set. These labels were recorded for subsequent comparison with the predicted labels produced by the trained machine learning model.

#### Machine Learning Model Selection and Fine Tuning

We utilized five machine learning models on the labelled training set —logistic regression, random forest, gradient boosting, support vector machine, and neural network. The dependent variable was pain sensitivity label (high/low) identified from the GMM and independent variables were sensorimotor PAF and ΔCME: the latter was typified as facilitator and depressor, depending on whether they showed an increase or decrease in map volume on Day 5 relative to Day 0, respectively. For each model, we identified optimized parameters through 5-fold cross-validation: we randomly divided the 80 participants into an internal training set of 64 participants (consisting of four equal folds of 16) and a validation set of 16. The optimized models in the internal training set were then employed to predict labels in the validation set to facilitate model selection. The model with the best performance (area under the curve) on the validation set was then locked in.

#### Test Set Prediction

The locked machine learning model was assessed on the test set. The participant IDs in this set did not coincide with those in the pain diary data, thereby preserving the double-blind nature of the analysis. By using the ground-truth labels (shuffled), predicted labels (unshuffled), and the shuffling order for the test set, we were able to evaluate the model’s performance by comparing the reordered predicted labels against the ground-truth labels established by the GMM. Performance was assessed via receiver operating characteristic (ROC) area under the curve (AUC), with 95% confidence intervals reported. AUC values between 0.7-0.8, 0.8-0.9 and 0.9-1 were considered “acceptable”, “excellent”, and “outstanding” respectively^29^.

## Results

### PAF/CME demonstrated good-excellent test-retest reliability

PAF and ΔCME showed good to excellent test-retest reliability across sessions (eFigures 5 and 7).

### Outstanding performance on the training validation set

Figure 2A shows the pain scores for participants in the training and test set classified as high and low pain sensitive. Figure 2B (upper) shows the performances of the models across the internal training and validation sets. Logistic regression was the winning classifier based on its outstanding performance (AUC=1.00[1.00-1.00]) when applied to the validation set (Figure 2B lower), with slower PAF and CME depression predicting higher pain with regression coefficients of -1.25 and -1.27 for PAF and CME respectively.

**Figure 2.**
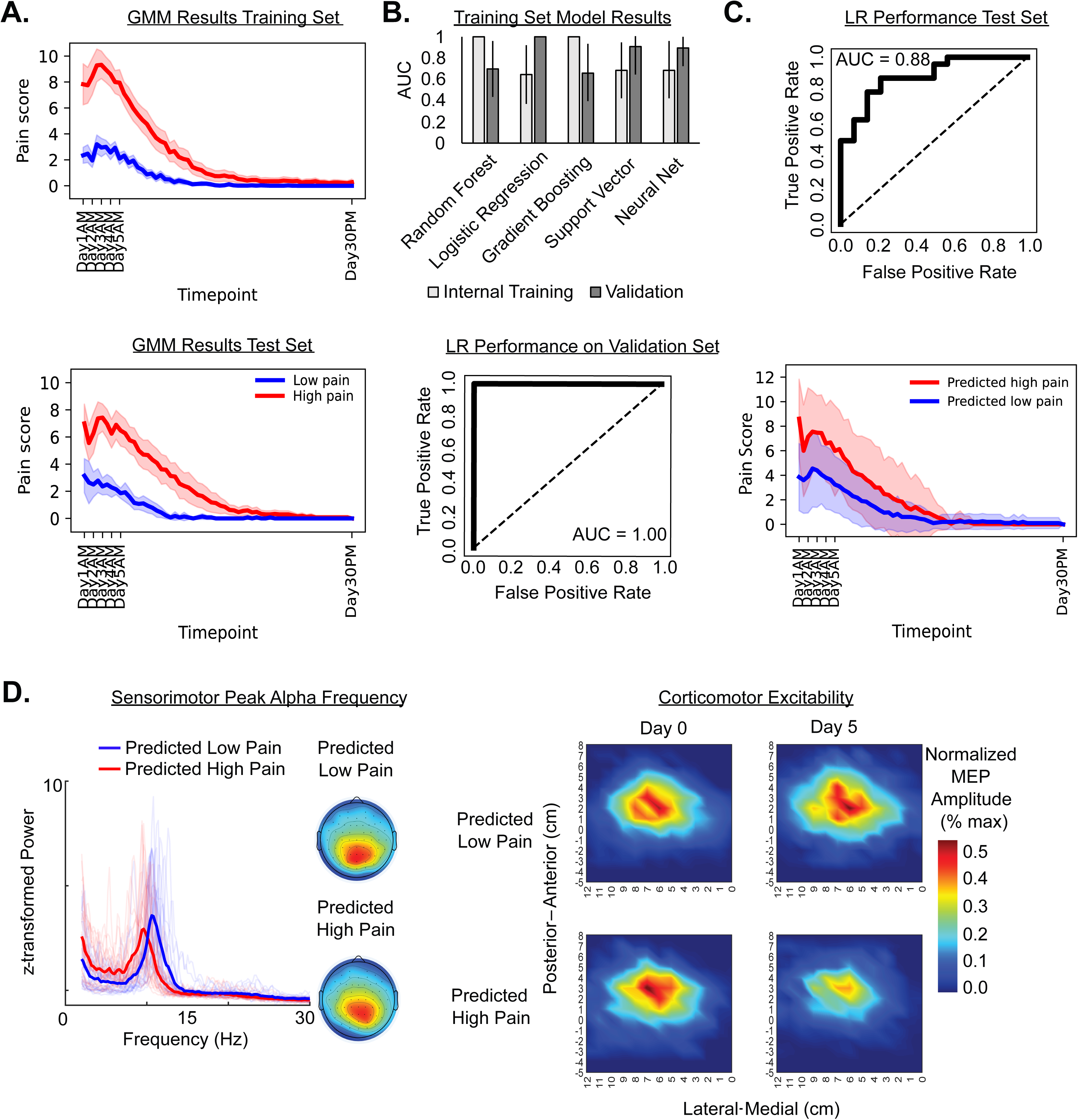
Performance of the combined PAF and CME biomarker on training and test set. **(A)** Results of the growth mixture modelling which categorized 80 participants in the training set (left) and 38 participants in the test (right) as high or low pain sensitive. Data shows mean pain score (chew + yawn pain rating) for each timepoint, while the shaded area shows 95% confidence intervals. **(B)** The upper panel shows performances (AUC [95% confidence intervals]) of various machine learning models for the internal training set and validation set. Logistic regression (LR) was chosen as the optimal classifier based on outstanding AUC of 100% as shown in the lower panel. **(C)** The upper panel shows the performance of the locked logistic regression model when applied to the test set, which was in the excellent range (AUC of 88%). The lower panel shows the pain trajectories (mean chew + yawn pain and 95% confidence intervals) of participants predicted to have high or low pain sensitivity based on the locked logistic regression model. **(D)** The left panel shows the individual and mean z-transformed spectral plots and topography of the sensorimotor alpha component on Day 0 for participants predicted to have high or pain sensitivity based on the locked logistic regression model. The right panel shows the mean motor cortex maps on Day 0 and Day 5 showing normalized motor evoked potential (MEP) amplitude (expressed as a proportion of the maximal MEP amplitude) for participants predicted to have high or low pain sensitivity based on the locked logistic regression model.

### Excellent performance on the test set

When the locked logistic regression model was applied to the test set, performance (Figure 2C upper) was excellent (AUC=0.88[0.78-0.99]). The optimal probability threshold for being classified as high pain sensitive was 0.40, with an associated sensitivity of 0.875 and specificity of 0.79. Applying this 0.40 probability threshold to our data, to be labelled as high pain sensitive, a facilitator would need a PAF<9.56, and a depressor would need a PAF<10.57. Figure 2C (lower) shows the differences in pain scores between participants predicted to have high or low pain. Visually one can observe slower PAF in those predicted to have high vs. low pain sensitivity (Figure 2D), This was statistically significant according to a two-sample t-test (*t*(48)=5.8, *p*<.001). Moreover, one can observe a decrease in CME within the masseter motor maps in those predicted to have high pain (Figure 2E), whereas those predicted to have low pain exhibited an increase in CME. The differences in ΔCME between these groups was statistically significant (*t*(48)=2.81, *p*=.007).

### A benefit for a *combined* signature

We reran the models to determine whether the combined PAF/CME signature out-performed each measure individually (eFigure 10). The performance of the PAF-only logistic regression model on the training validation and test set were respectively excellent (AUC=0.95[0.84-1.00]) and outstanding (AUC=0.83[0.70-0.96]). The performance of the CME-only logistic regression model for the training validation and test set were respectively excellent (AUC=0.88[0.69-1.00]) and acceptable (AUC=0.75[0.60-0.91]).

### Inclusion of covariates did not improve model performance

We evaluated the performance of the biomarker combined with covariates. As there were many variables, we applied feature selection, i.e. filtering features by inspecting p-values when associating predictors and labels, and using parameter tuning to optimize the coefficients associated with the filtered features. Five features were subsequently selected and optimized – Sensorimotor PAF, CME, Sex, Pain Catastrophizing Scale (PCS) Total and PCS Helplessness. The associations between labels and biomarkers/covariates in the training vs. test set, and performance of the models are shown in Figure 3A and 3B. When including these five features, the performance of the logistic regression model (regression coefficients of -0.86, -0.69, 0.64, 0.02 and 0.06 for PAF, CME, Sex, PCS Total and PCS Helplessness respectively) was outstanding (AUC=1.00[1.00-1.00]) and excellent (AUC=0.81[0.67-0.95]) for the training validation and test set were respectively. Thus, the model with biomarkers only outperformed the model including covariates.

**Figure 3.**
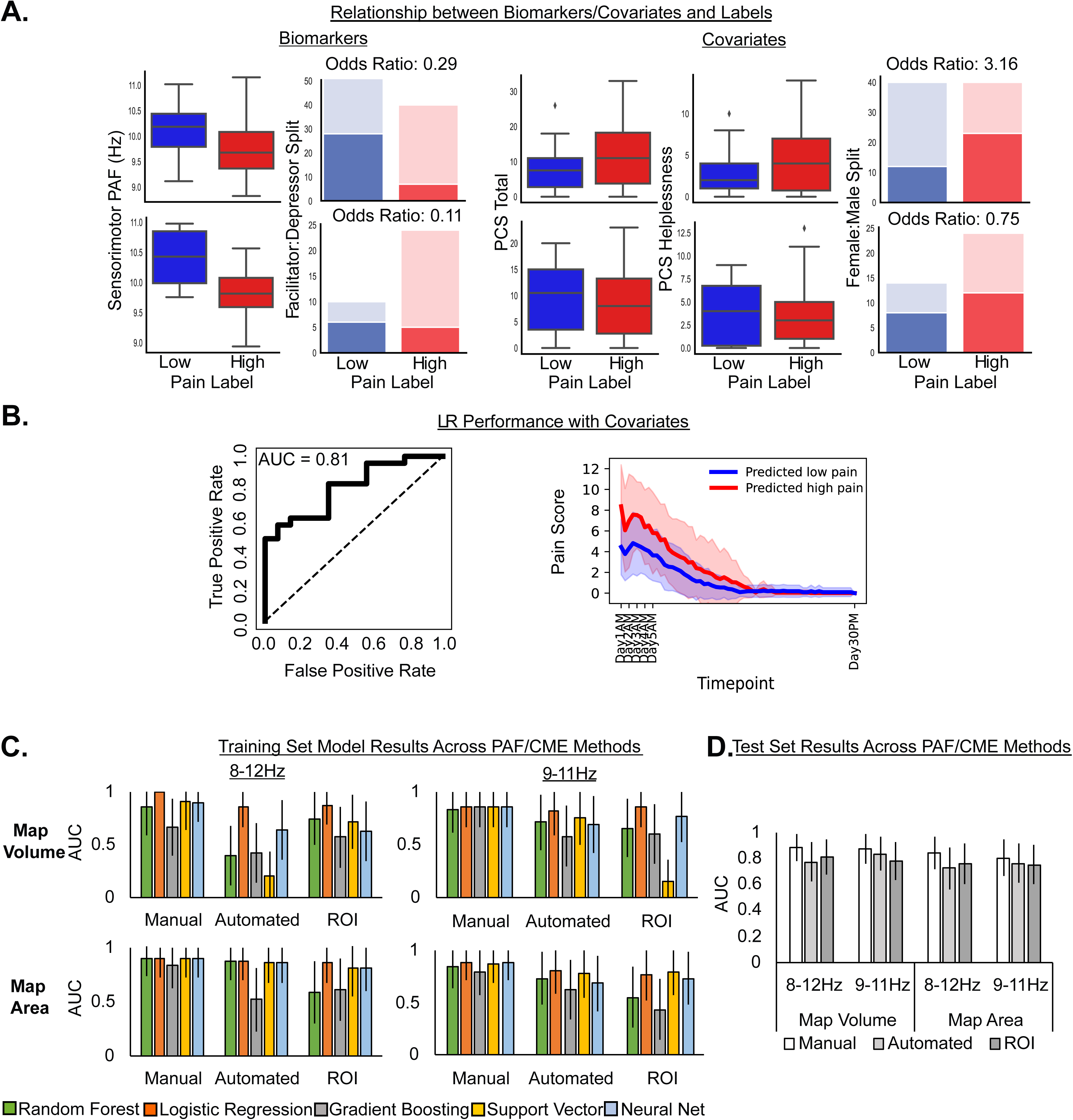
Performance of the combined PAF and CME biomarker on training and test set when including covariates, and across PAF/CME calculation methods. **(A)** Visualisation of biomarkers and covariates for the training and test sets across high (red) and low (blue) pain labels identified from the GMM. Data on PAF, PCS total and PCS helplessness are plotted as boxplots, while data on CME and Sex are plotted according to facilitator: depressor and female: male split respectively, including odd ratios. A lower odds ratio means a lower probability of high pain sensitive individuals belonging to the facilitator or female categories. For PAF and CME, low pain was associated with fast PAF and CME facilitation for both training and test sets. In contrast, the relationship between covariates and labels were in the opposite direction for the training and test set, suggesting the relationship between biomarkers and labels was consistent. **(B)** The left panel shows the performance of the locked logistic regression model on the test set when including covariates in the model. The right panel shows pain trajectories (mean chew + yawn score and 95% confidence intervals) of participants predicted to have high or low pain sensitivity based on the locked logistic regression model including covariates. **(C)** The performance of each machine learning model (AUC [95% confidence intervals]) on the training validation set across different PAF/CME calculation methods. This includes the sensorimotor component chosen manually after an independent component analysis, component identified using an automated script after an independent component analysis, or using a sensorimotor region of interest (ROI, mean of Cz, C3 and C4) in electrode space, to calculate PAF. We also looked at different frequency windows for computing PAF (8-12Hz vs. 9-11Hz) or CME calculated using map area or map volume. **(D)** The performance of the locked logistic regression model (AUC [95% confidence intervals]) when applied to the test set, across different PAF/CME calculation methods.

### Results were reproducible across methodological choices

To determine whether our results were robust across different methodological choices, we repeated the analysis using different PAF calculation methods, including component level data (with the sensorimotor component chosen manually or using an automated script) vs. sensor level data (with a sensorimotor region of interest), using different frequency windows (8-12Hz vs. 9-11Hz) and using different CME calculation methods (map volume vs. map area). We found that, regardless of the choices, logistic regression was the best or equal-best performing model when applied to the training-validation set (Figure 3C), with AUCs varying from acceptable (AUC=0.77) to outstanding (AUC=1.00). When the locked models were applied to the test set, performance varied from acceptable (AUC=0.73) to excellent (AUC=0.88) (Figure 3D). Lastly, excellent performance was demonstrated when the data was analysed two other ways (eFigure 11 and 12): where GMM pain labels were established using the whole 30 days rather than the first 7 days (training validation AUC=0.84[0.64-1], test AUC =0.89[0.79-0.99]), and when missing pain diary data was not imputed (training validation AUC=0.81[0.6-1], test AUC=0.89[0.79-0.99]).

## Discussion

A full-scale analytical validation of the PAF and CME biomarker signature was conducted using a prolonged pain model. In an initial training set (n=100), we found that a logistic regression was the optimal classifier based on its outstanding performance (AUC=100%), with slower PAF and CME depression predicting higher pain. When this model was applied to an independent test set, the AUC was excellent (AUC=88%). PAF/CME showed good-excellent test-retest reliability, and results were reproduced across a range of methodological parameters. Inclusion of covariates did not improve model performance, suggesting the model including biomarkers only was more robust. Overall, the combination of sample size, pain model validity, and biomarker accuracy, reproducibility and reliability suggest the PAF/CME biomarker signature has substantial potential for clinical translation.

Our results suggest that individuals who have slow PAF prior to an anticipated prolonged pain episode and show corticomotor depression during a prolonged pain episode, are more likely to experience higher pain. Model performance was higher combining the two, suggesting consideration of both ascending sensory and descending motor pain processing mechanisms provides more information regarding pain sensitivity. Note that our study used a cohort of healthy participants with strict inclusion/exclusion criteria and an experimental pain model. While this may limit generalizability to clinical populations, the use of a standardized sample/design is a requirement of pre-clinical analytical validation, and an essential first step in the discovery pipeline toward a clinical biomarker signature. Moreover, there is already evidence that the proposed biomarker is generalizable to clinical contexts. A recent study showed that individuals with slower PAF experienced more pain following a thoracotomy^20^. Furthermore, individuals with lower CME during the acute stages of low back pain were more likely to develop chronic pain at 6-months follow-up^35^. This suggests PAF and CME shows promise in being used in pre-operative and/or post-operative/post-injury contexts to classify high or low pain sensitive individuals. Given that higher acute pain predicts the development of chronic pain^24^ PAF and CME could potentially be used as susceptibility biomarkers for the transition from acute to chronic pain.

There are several aspects of our study which stand out. The first is sample size: with recent advancements in machine learning, it has become possible to conduct analytical validation of pain biomarkers. However, deep learning requires a large amount of labelled samples to conduct rigorous training on validation and test sets^8^. Unfortunately, many pain susceptibility biomarker studies have not been sufficiently sampled to adopt such approaches^9,10^, and the ones that have used machine learning failed to reach the sample sizes similar to that of the present study^1,2^.

Another strength of our findings is reproducibility. Previous work has shown similar associations between slower PAF and higher upper limb pain, post-operative thoracic pain and chronic pain in various body regions^17,19,20^ as well as CME depression and higher upper limb pain, chronic patellofemoral pain and development of chronic low back pain^22,23,36,37^. The present study replicated these results in a model of jaw pain, suggesting the biomarker signature may be generalizable to pain more broadly. Note that some studies have not shown a negative relationship between PAF and pain sensitivity^38,39^ or a positive relationship between CME depression and pain sensitivity^31^. However, these studies were not sufficiently sampled to conduct analytical validation of the kind presented in this study. Nonetheless, the mixed findings could also arise from differences in methodological choices in the estimation of PAF e.g. frequency windows^39^ and use of sensor vs. component space data^40^ and estimation of CME e.g. map volume^22^ vs. area^31^. For this reason, we repeated the main analysis using different methodological choices and found at least acceptable AUCs, supporting the reproducibility of our results.

The PAF/CME measures demonstrated good-excellent reliability. Reliability is a desirable characteristic which assists in the widespread application of pain biomarkers^8^. We found that participants exhibit stable PAF across days despite the presence of pain, and even when considering different methodological factors that may influence the reliability such as pre-processing pipeline, recording length and frequency window^14^. Indeed, reliable PAF was found with a recording length as short as 2 minutes and minimal data pre-processing. We also showed that those who show CME depression on Day 2 are also likely to show CME depression on Day 5 (and vice versa for those who show CME facilitation). This was shown even when an automated method of determining MEP amplitude on each trial was applied. Thus, our work not only shows that PAF and CME can predict pain, but the relative ease with which reliable PAF/CME data can be obtained is promising for subsequent clinical translation.

Another strength of this study is our pain model. While other pain biomarker studies have shown promising results, these studies were restricted to pain models utilizing transient painful stimuli lasting seconds to minutes^11-13^. The brief nature of the painful stimuli questions the external validity of these findings and limits generalizability to clinical populations. In contrast, the present study used a prolonged pain model lasting weeks. Several other studies have shown that injections of NGF to the neck, elbow or masseter muscles can mimic symptoms of clinical neck pain^41^, chronic lateral epicondylalgia^27^ and TMD^28^ respectively. Thus, the observed relationships between PAF/CME and pain in the present study show promise in terms of clinical applicability.

Lastly, the PAF/CME biomarker demonstrated high performance. A previous study found that connectivity between medial prefrontal cortex and nucleus accumbens in 39 sub-acute low back pain patients (pain duration 6-12 weeks) could predict future pain persistence at ∼7, 29 and 54 weeks, with AUCs of 67-83%^1^. Another study on 24 sub-acute low back pain patients showed that white matter fractional anisotropy measures in the superior longitudinal fasciculus and internal capsule predicted pain persistence over the next year, with an AUC of 81%^2^. Though the present did not directly assess the transition to chronic pain, our AUCs of 100% (validation set) and 88% (test set) appear comparatively high. We therefore encourage future clinical studies to determine whether PAF/CME can predict the transition from acute to chronic pain.

## Conclusions

A novel biomarker signature comprised of PAF and CME accurately and reliably distinguishes high and low pain sensitive individuals during prolonged jaw pain with an excellent AUC of 88% in an independent test set. No other pain biomarker study has shown this combination of biomarker accuracy, reproducibility, reliability and pain model validity, suggesting high potential for clinical translation, particularly in predicting the transition from acute to chronic pain.

## Data Availability

The corresponding author had full access to all the data in the study and takes responsibility for the integrity of the data and accuracy of the data analysis. Code and relevant diary, questionnaire, and demographic data are available at https://github.com/DrNahianC/PREDICT_Scripts. The raw data supporting the findings of this study are available on OpenNeuro (https://openneuro.org/datasets/ds005486/versions/1.0.0) for the EEG data and OSF (https://osf.io/r3m9g/) for the TMS data.

https://github.com/DrNahianC/PREDICT_Scripts

https://osf.io/r3m9g/

## Acknowledgements

This project was funded by the National Institutes of Health (R61 NS113269/NIH/NINDS HHS/United States). Clinical Trial Registration: NCT04241562 https://classic.clinicaltrials.gov/ct2/show/results/NCT04241562?view=results. The authors have no conflicts to declare. The corresponding author had full access to all the data in the study and takes responsibility for the integrity of the data and accuracy of the data analysis. Code and relevant diary, questionnaire, and demographic data are available at https://github.com/DrNahianC/PREDICT_Scripts. The raw data supporting the findings of this study are available on OpenNeuro (https://openneuro.org/datasets/ds005486/versions/1.0.0) for the EEG data and OSF (https://osf.io/r3m9g/) for the TMS data.

