## Supplementary Material for "A novel cortical biomarker signature predicts individual pain sensitivity"

**Online-Only Supplements- Table of Contents**

| eMethods | 2 |
| --- | --- |
| Sample Size Justification  Inclusion/Exclusion Criterion | 2 |
|  | 2 |
| Participant Exclusions | 2 |
| Data Collection Procedures | 2 |
| EEG Data Processing | 4 |
| eFigure 1 – Pipeline for Automated Component Selection | 5 |
| eFigure 2 – Template for Automated Component Selection | 5 |
| TMS Data Processing | 5 |
| Pain Diary Data Processing | 6 |
| eResults | 6 |
| Questionnaire and Demographic Data | 6 |
| eTable 1 – Baseline Questionnaire Results | 7 |
| eTable 2 - Race demographics | 7 |
| Pain Diary Data | 8 |
| eFigure 3 – Means for Pain Diaries | 8 |
| Pressure Pain Threshold Data | 8 |
| eFigure 4 – Means for Pressure Pain Thresholds | 8 |
| Additional EEG Analysis | 9 |
| eFigure 5 – PAF Test-Retest Reliability | 9 |
| Additional TMS Analysis | 10 |
| eFigure 6 – TMS Motor Maps | 10 |
| eFigure 7 – CME Test-Retest Reliability | 11 |
| Pain Diary Separated by PAF/CME Classifications | 11 |
| eFigure 8 – Pain Diary Separated by PAF/CME Classifications | 11 |
| eFigure 9 – Comparison of pain scores for fast vs. slow PAF and facilitator vs. depressor | 12 |
| Additional Modelling Results | 12 |
| eFigure 10 – Model Performance of PAF/CME markers alone | 12 |
| eFigure 11 – Model Performance without linear interpolation of pain data  eFigure 12 – Model Performance when using the Days 1-30 for pain labelling | 13 |
|  | 13 |
| eFigure 13 – Model performance when including CME as a continuous variable | 14 |
| References | 14 |

**eMethods**

**Sample Size Justification**

Preliminary data indicates strong associations between pain severity and PAF/ΔCME [1-5]. The design of the current discovery-based study is not amenable to traditional p-value based power calculations, but rather is predictive. In machine learning studies, training set classification and test set accuracy benefits from larger sample sizes. We have chosen a sample size of 150 participants, which provides confidence in the resultant classification and accuracy estimates and is considerably higher than other biomarker validation studies [6].

**Inclusion/Exclusion Criteria**

An age inclusion criterion of 18-44 was chosen based on the orofacial pain prospective evaluation and risk assessment (OPPERA) prospective cohort study that demonstrated an incidence rate of first onset temporomandibular disorder (TMD) of 2.5% per annum among 18 to 24-year olds and 4.5% per annum among 35 to 44-year olds [7]. Participants were excluded if they presented with any acute pain, had a history of any chronic pain, any major medical complaints, psychiatric or salivary gland conditions, were pregnant and/or lactating, exhibited frequent alcohol, opioid or illicit drug use in the past 3 months, or were contraindicated for TMS (e.g., metal implants, epilepsy) as assessed using the Transcranial Magnetic Stimulation Adult Safety Screen questionnaire [8].

**Participant Exclusions**

Participants were recruited between November 2020 and October 2022 through notices posted online, at universities across Sydney, Australia or by contacting participants held on a database at NeuRA. Nine participants did not complete both nerve growth factor (NGF) injections (withdrew during the first session before the injection or before attending the second) and were excluded from all analyses. An additional 7 participants completed both NGF injections but had missing electroencephalography (EEG) and/or transcranial magnetic stimulation (TMS) data on Day 5. These participants were included in the analyses as they still completed the electronic pain diaries and could still be classified as a facilitator or depressor using their Day 2 TMS data based on previous research [9]. This left 150 participants in the final sample. Participants were recruited via advertisements placed on community notice boards, social media platforms and a healthy participant volunteer database.

**Data Collection Procedures**

**Baseline Questionnaires.** On Day 0 prior to pressure pain threshold, peak alpha frequency (PAF) and corticomotor excitability (CME) measurements, participants completed questionnaires pertaining to sex, race, physical and mental health, and experiences with pain, to be included as covariates in the predictive modelling. These include: i) Pain Catastrophizing Scale [10], ii) Perceived Stress Scale [11], iii) Sleep Scale, iv) Patient health questionnaire [12], v) Generalized Anxiety Questionnaire [13], vi) Tobacco Alcohol Prescription Medication and Other Substances [14], vii) Pennebaker Inventory of Limbic Languidness Questionnaire [15], viii) Short-Form-8 Health Questionnaire [16] and ix) The Brief Pain Inventory Pain Severity and 7-item Interference subscales [17, 18].

**Pressure Pain Thresholds.** Prior to PAF and CME measurement on Day 0, 2 and 5, pressure pain thresholds (PPTs) were assessed at 5 sites: i) the right masseter muscle, ii) the right temporalis muscle, iii) the right temporomandibular joint, iv) the right trapezius muscle and v) the right thumbnail. A 1-minute rest was observed between one round of the 5 measurements and 3 measures were made at each site. Pressure was applied at a rate of 40 kPa/s using a handheld algometer (Somedic, Horby, Sweden) perpendicular to the surface of the skin. Participants were asked to push a button when the sensation of pressure first changed to one of pain. The position of each site was recorded with a tape measure to ensure consistency of algometer placement across testing days. The average of the 3 trials at each site was used for analysis.

**Peak Alpha Frequency.** Participants were seated in a comfortable chair. Scalp EEG was recorded using the Brain Products platform (BrainVision Recorder, Vers. 1.22.0101 with actiCHamp Plus, Brain Products GmbH, Gilching, Germany). Recording was initially done at a sampling rate of 25000 Hz (participants 1–16), though this was subsequently reduced to 5000 Hz (participants 17–159) as the higher sampling rate was deemed unnecessary to obtain reliable resting state alpha activity. Signals were recorded from 63 active electrodes (actiCAP slim, Brain Products GmbH, Germany), embedded in an elastic cap (EASYCAP, EASYCAP GmbH) in line with the 10–10 system. Recordings were referenced online to ‘FCz’ and the ground electrode placed on ‘FPz’. Electrode impedances were maintained below 25 kOhms during setup. Based on previous research ([19, 20]) and the actiCHamp manual, we anticipated negligible signal loss with this impedance cut-off. Once setup was complete, the lights were switched off with ambient noise reduced to a minimum. Participants were instructed to relax their muscles and keep their eyes closed while remaining awake. The resting-state EEG signal was then recorded for 5 min.

**Corticomotor Excitability.** Participants were seated in a comfortable chair and viewed a monitor that displayed electromyographic (EMG) signals used for live feedback. Bipolar surface electrodes were used to record EMG from the right masseter muscle. A belly-tendon montage was used, with the active (belly) and reference (tendon) electrodes placed along the mandibular angle, and ground electrode placed on the right acromion. EMG signals were amplified (x 1000) and filtered (16 to 1000Hz), and digitally sampled at 2000 Hz.

It is common to assess CME from the masseter muscle during active contraction that is at a certain percentage of the maximum voluntary contraction (MVC) (see [21, 22]). This is because it is challenging to obtain reliable MEPs from the masseter muscle at rest [23, 24]. We instructed participants to contract at 20% MVC as this provided a balance between obtaining sufficiently large MEPs for analyses [25], while minimizing fatigue. To measure MVC, participants were actively encouraged to clench their jaw as hard as possible for 3 seconds on 3 separate trials, with a 1-minute rest break between trials. The MVC was computed based on the average of the 3 trials, and 20% MVC was computed based on this average. Participants were provided with practise at maintaining a masseter contraction of 20% MVC by receiving live feedback from the EMG signal.

Single, monophasic stimuli were delivered using a Magstim unit (Magstim Ltd., UK) and 70mm figure-of-eight flat coil. In line with previous studies investigating optimal coil orientation for inducing masseter MEPs, an angle of 90 degrees between the anterior-posterior line and the coil handle was used [26]. This orientation induced a current in the lateral-to-medial direction. Participants wore a swim cap with a grid of 1cm x 1cm resolution. The scalp site evoking the largest MEP (i.e., the “hotspot”) during active contraction was then determined.

The TMS motor threshold assessment tool [27] was used to determine the active motor threshold (AMT). This uses maximum likelihood parametric estimation by sequential testing to compute the minimum intensity required to evoke an MEP with a 50% probability. A reliable MEP was identified if the EMG waveform between ~5-15ms [28, 29] after the TMS pulse was visually larger in amplitude relative to background contraction.

The procedure for mapping has been described in detail previously [30, 31]. The TMS intensity was set at 120% AMT. During active contraction, 3 stimuli were delivered at each location around the grid, starting at the hotspot. The number of stimulation sites was pseudorandomly increased until an MEP was no longer observed (i.e., no reliable MEP in all 3 trials at all border sites). Note that on Day 2 and 5, the mapping procedure began immediately after the MVC was determined, with the motor hotspot and threshold procedures skipped. In line with the approach used in previous studies [32-34], the hotspot location and test stimulus intensity determined on Day 0 was used for subsequent mapping procedures on Day 2 and Day 5. This approach ensures that any changes in motor cortical maps occurring as a result of NGF-injection can be observed. Although we did not re-assess AMT on Day 2 and 5, a previous study showed no change in AMT in the masseter after NGF injection [22].

**Nerve Growth Factor.** The Injection of Recombinant Human β-NGF is used in several labs worldwide [35, 36] for the induction of muscle soreness in healthy individuals, and while invasive and painful, is safe and is associated with minimal adverse effects[37]. The NGF injections were provided at the end of the Day 0 and Day 2 sessions. To prepare the masseter muscle for injection, the surface was cleaned using alcohol wipes. A sterile solution of NGF (dose of 5 μg [0.2 ml]) was then administered as a bolus injection into the muscle belly of the right masseter using a 1-ml syringe with a disposable needle (27-G). The needle was inserted perpendicular to the masseter body until bony contact was reached, retracted ∼2 mm, and NGF injected.

**Electronic Pain Diaries.** Participants completed electronic pain diaries using a computer, tablet or phone at 10 am and 7 pm each day from day 0 to 30. Within the electronic diary, participants were asked to rate their pain intensity on an 11-point numerical rating scale anchored with ‘no pain’ at zero and ‘worst pain imaginable’ at 10 at rest and during chewing, swallowing, drinking, talking, yawning and smiling.

**EEG Data Processing**

**Main EEG Pipeline – Manual Sensorimotor Alpha Component Selection.** EEG data processing was conducted using custom MATLAB (R2020b, The Mathworks, USA) scripts implementing the EEGLAB (eeglab2019_1) [38] and Fieldtrip (v20200215) toolboxes [39]. The pre-processing pipeline matched those used in previous studies investigating the relationship between PAF and pain [40]: data downsampled to 500 Hz, re-referenced to common average, and band-pass filtered between 2 and 100 Hz using an FIR filter. Channel data was visually inspected, and overtly noisy channels were removed and the signal re-referenced. Data were segmented into 5 s epochs, and epochs containing marked muscular artefacts were manually rejected. Independent component analysis was applied to the data. We visually inspected the frequency-spectra of the components, and identified components that had a clear alpha peak (8–12 Hz) and a scalp topography suggestive of a source predominately over the sensorimotor cortices [40].

The power spectral density of the chosen component was derived in 0.2 Hz bins. The 2–50 Hz range was extracted using Fast Fourier Transform. A Hanning taper was applied to the data to reduce edge artefacts. PAF was calculated using the Centre of Gravity method, using the following equations:

$$CoG=\frac{\sum_{i=1}^{n} f_{i}a_{i}}{\sum_{i=1}^{n} a_{i}}$$

Where f_i_ is the ith frequency bin within the frequency window of interest, and a_i_ represents the spectral amplitude at f_i_.

**Automated Sensorimotor Alpha Component Selection Method.** As in the pipeline presented in the main paper, this pipeline was identical up the point where ICA was applied. All components which displayed a clear peak in the alpha band (8 to 12 Hz) were compared to a sensorimotor spatial template generated from findings of prior work [40] (eFigure 1). Specifically, power within the alpha band at each sensor in our data was correlated to the power within the alpha band at each sensor of the template.

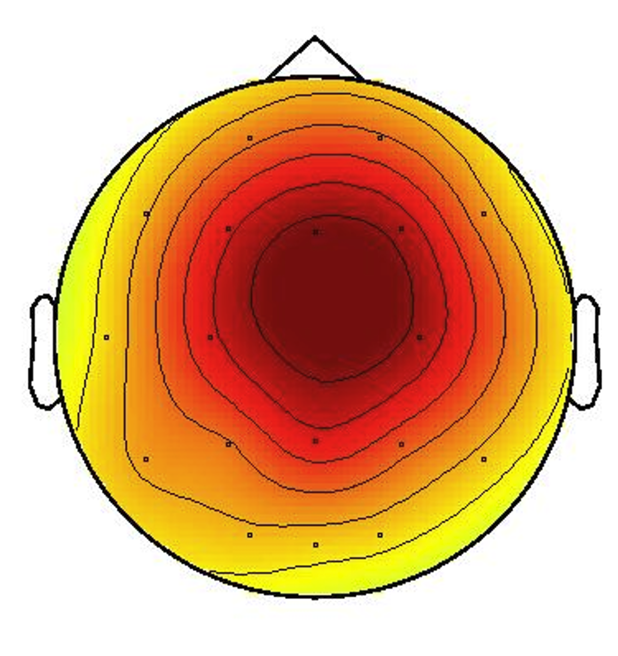

*eFigure 1.* Alpha band power of each ICA component was compared against a sensorimotor template generated from previous work [40].

The component which displayed both a spectral peak in the alpha band and that displayed the strongest spatial correlation (calculated via Spearman’s Rank method) to the template was selected as representing sensorimotor alpha activity.  Components were selected based on the absolute value of the correlation. Spatial distribution of alpha activity was considered regardless of polarity of the correlation strength. Given outputs of ICA may differ upon each run, this process was iterated ten times per participant to improve the reliability of the automation, resulting in ten selected components for each participant. Components with the highest correlation to the template was selected for PAF calculation. Once the best-fitting component was selected, PAF was calculated using the centre of gravity method, resulting in a single PAF value for each participant. See eFigure 2 for details of the pipeline.

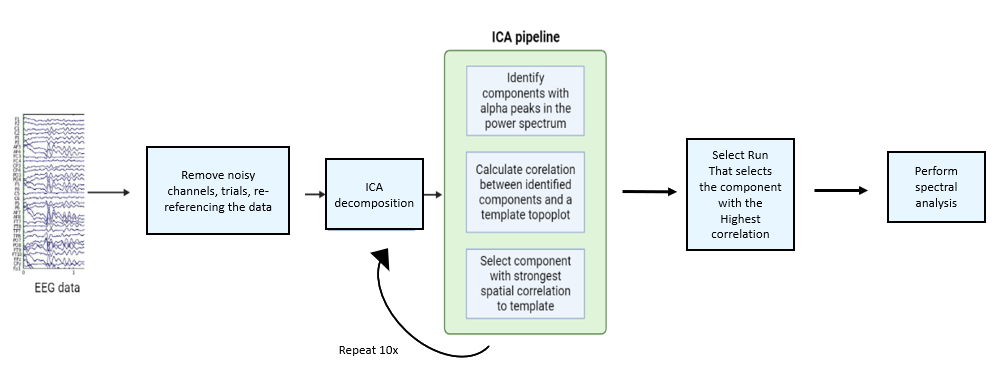

eFigure 2. Pipeline for automated selection of sensorimotor alpha component.

**Sensor Level PAF Calculation.** This pipeline was identical to that used in our previous work [9]. As in the pipeline presented in the main paper, this pipeline was identical up the point where ICA was applied – however ICA was not applied to identify a sensorimotor alpha component. Rather, ICA was applied to remove eyeblink and saccade artefacts from the sensor space data. Once this was done, PAF was calculated using the centre of gravity method as described in the main paper, and averaged across sensorimotor region of interest (Cz, C3, C4).

**TMS Data Processing**

**Corticomotor Excitability.** TMS data was processed using a custom MATLAB script. The MEP onsets and offsets across were fixed to 7.3 and 15.3 ms specifically based on our previous study which showed that using a fixed MEP window is just as reliable as manually choosing the offsets and onsets[9]. The RMS of background EMG was determined using a fixed window between 55 and 5ms before the TMS pulse [41]. This was subtracted from the RMS of the MEP window to determine MEP amplitude. The mean MEP amplitude at each stimulation site was determined. Map volume was calculated by summing the amplitudes of sites exhibiting >10% of the maximum MEP amplitude [42]. Corticomotor excitability (CME) was computed by determining the difference between the map volumes on day 0 and day 5 (Day 5 – Day 0). In this framework, positive and negative CMEs were typified as facilitator and depressor respectively, labelled as 1 and 0. Any missing values encountered in this data subset were addressed through multivariate imputation.

**Additional Parameters**

***Map Area*.** Whereas map volume was calculated by summing the amplitudes of sites exhibiting >10% of the maximum MEP amplitude, map area was defined as the number of sites (measured in cm^2^) on the grid which exhibited an MEP amplitude that was greater than 10% of the maximum MEP amplitude.

***Map Centre of Gravity*.** The centre of gravity (CoG), defined as the amplitude-weighted centre of the map in either the anterior-posterior direction or the medial-lateral direction, was calculated, using the following formula: (CoG=${\sum V_{i}xX_{i}}/{\sum V_{i}}$, ${\sum V_{i}xY_{i}}/{\sum V_{i}}$; *V_i_*=mean MEP amplitude at each site with the coordinates *X_i_, Y_i_* ) [43, 44].

**Pain Diary Data Processing**

We consolidated the morning and afternoon recordings into a single table to maintain a chronological order of pain intensity records, spanning from Day 1 through Day 30. Any missing values within this data were addressed via multivariate imputation.

**eResults**

**Questionnaire Data**

eTable 1 shows means and SDs for each of the baseline questionnaires.

*eTable 1. Scores on baseline questionnaires. PCS = Pain Catastrophizing Scale, BPI = Brief Pain inventory, SF8 = Short-Form 8 Health Questionnaire, SS = Sleep Scale, PHQ = Patient Health Questionnaire, GAD = Generalised Anxiety Disorder, TAPS = Tobacco Alcohol Prescription Medication and Other Substances, PSS = Perceived Stress Scale, PILL* = *Pennebaker Inventory of Limbic Languidness Questionnaire.*

| Questionnaires | Score |
| --- | --- |
| PCS Rumination | 3.8±3.2 |
| PCS Magnification | 2.1±2.1 |
| PCS Helplessness | 3.5±3.3 |
| PCS Total | 9.3±7.5 |
| BPI Severity | 0.1±0.4 |
| BPI Interference | 0.5±1.1 |
| SF8 PCS | 1.8±0.6 |
| SF8 MCS | 1.7±0.6 |
| SS Time total | 2.2±3.7 |
| SS Total | 45.4±5.2 |
| PHQ Total | 0.7±1.1 |
| GAD2 Total | 0.8±1.0 |
| TAPS | 0.8±1.5 |
| PSS Total | 18.6±8.3 |
| PILL Total | 86.4±20.9 |

eTable 2 shows the number of participants in each race category

*eTable 2. Number of participants in each race category*

| Race | Number of participants |
| --- | --- |
| Asian | 66 |
| White | 59 |
| Unknown | 24 |
| More than one race | 9 |
| American Indian | 0 |
| Black | 1 |
| Hawaiian | 0 |

**Pain Diary Data**

eFigure 3 shows the mean ratings across time for each of the pain diary items. Scores for pain upon chewing and yawning were higher than for all other pain measures, suggesting these two measures were the most relevant to the TMD NGF Model.

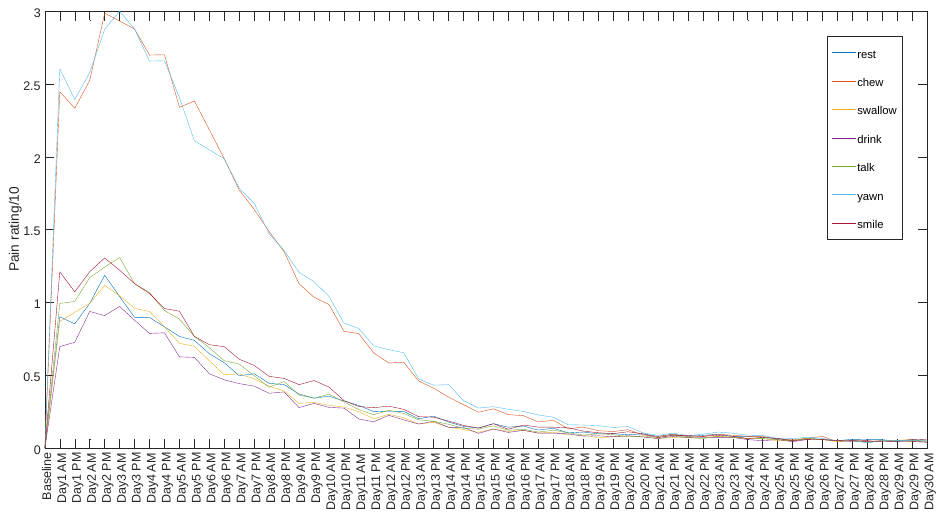

*eFigure 3.* Mean scores for pain at rest, chewing, yawning, swallowing, drinking, talking and smiling.

**Pressure Pain Thresholds Data**

eFigure 4 shows the mean PPTs across time for each location. Our previous analyses [45] has shown that the reduction in PPT was larger in the masseter compared to any other location.

Pressure Pain Threshold (kPA)

|  | Day0 | Day2 | Day5 |
| --- | --- | --- | --- |
| masseter | 161.841 | 60.5511 | 48.2391 |
| temporalis | 234.229 | 210.349 | 223.961 |
| trapezius | 325.683 | 301.996 | 302.552 |
| TMJ | 167.618 | 156.844 | 165.343 |
| thumb | 323.285 | 310.753 | 316.092 |

*eFigure 4.* Mean (± SEM) PPTs across time for each location

**Additional EEG Analysis**

**Reliability of Sensor level PAF.** eFigure 5 shows data pertaining to the reliability of the sensor level PAF data across days. Similar to our previously published data [9] which consistent of half of the participants in the present data set , we found that regardless of whether standard pre-processing was used, the length of the recording window, or the choice of frequency window for PAF, the ICC of sensor level PAF was excellent across days (green represents an ICC > 0.8).

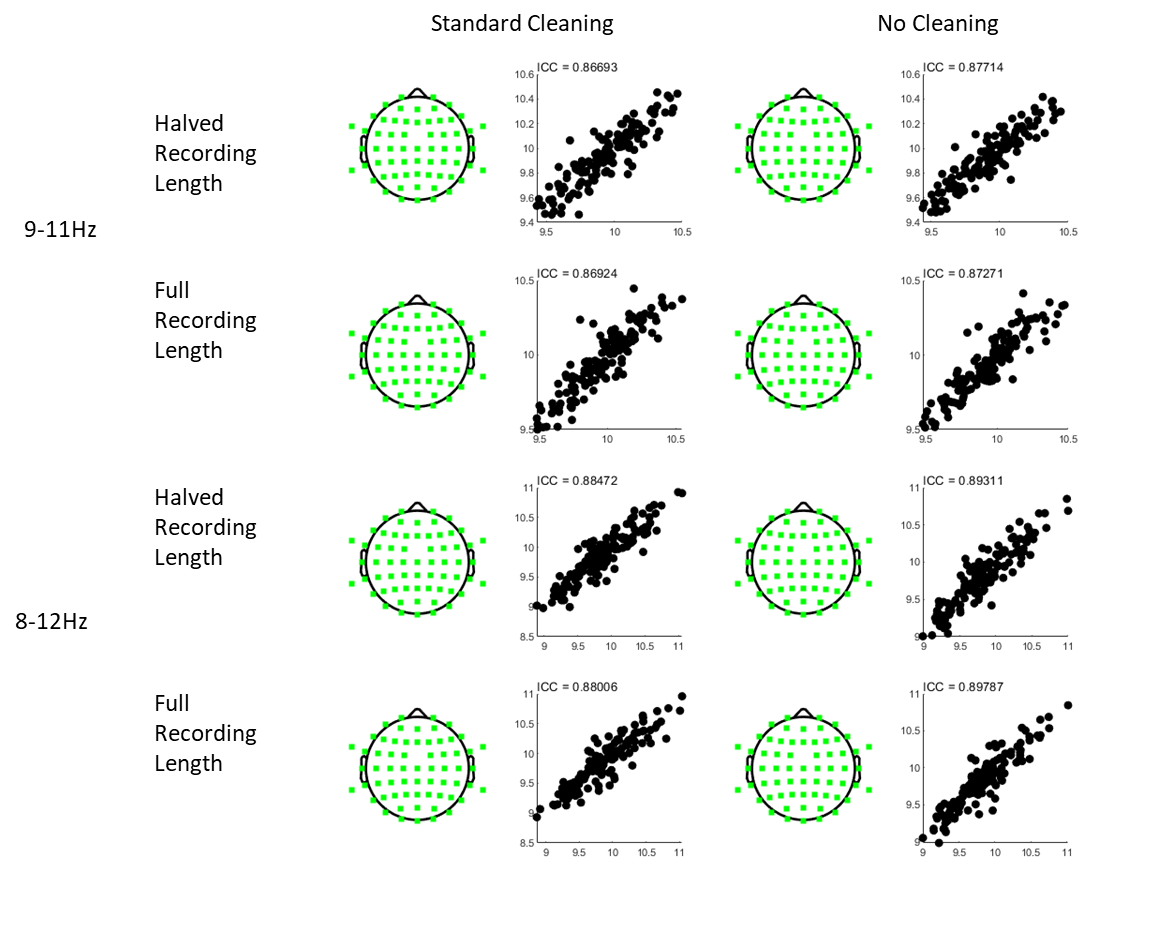

*eFigure 5.* Topographies and scatterplots of intraclass coefficients (ICCs) for peak alpha frequency (PAF) as a function of pre-processing pipeline, recording length and frequency window and calculation method. Green colours represents excellent test-retest reliability (ICC>0.8) of peak alpha frequency (PAF) in a channel between sessions. The scatterplots show PAF on Day 0 plotted against the mean PAF of Day 2 and Day 5 for a representative channel (Cz)

**Performance of the Automated Sensorimotor Component Selection Method.** In 9/150 participants, the automated script could not identify a component with a clear alpha peak. For these participants, components with potential alpha peaks were identified manually. The correlations between the chosen component and the sensorimotor template varied from 0.69 to 1, and the mean correlation was 0.85. PAF calculated using automated component had a correlation of 0.67 when compared to manual component selection, and 0.85 when compared to sensor level PAF. This data suggests the automated component selection method generated good-excellent consistency with the template, and good-excellent consistency with other PAF calculation methods.

**Additional TMS Analysis**

**Characterisation of Motor Maps.** eFigure 6 shows the mean normalized motor maps across all participants. The mean non-normalized map volume (in mV) was 0.45 ± 0.41 on Day 0, 0.42 ± 0.44 on Day 2 and 0.39 ± 0.44 on Day 5. The mean centre of gravity in the anterior-posterior dimension was 2.63cm ± 0.80 on Day 0, 2.44cm ± 1.0 on Day 2, and 2.54cm ± 0.82 on Day 5. The mean centre of gravity relative to the vertex in the medial-lateral dimension (in cm) was 6.72cm ± 0.93 on Day 0, 6.69cm ± 0.99 on Day 2, and 6.54cm ± 0.94 on Day 5. The mean map area was 13.95cm^2^ ± 5.15 on Day 0, 14.45cm^2^ ± 5.84 on Day 2, and 13.93cm^2^ ± 5.96 on Day 5.

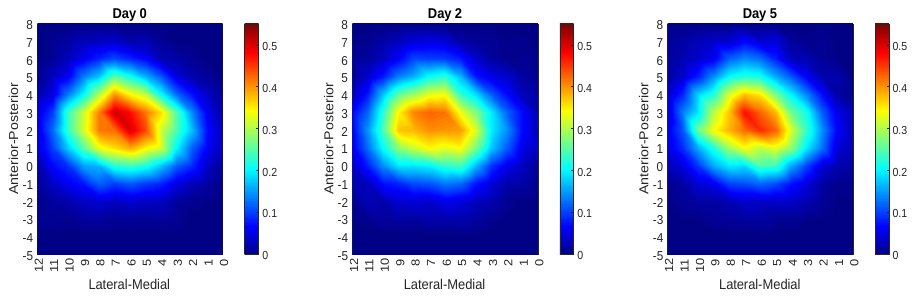

*eFigure 6.* Mean (normalized) motor maps across each day, showing the mean MEP amplitude at each location on the grid (expressed as a proportion to the highest MEP amplitude for each map).

eFigure 7 shows the correlation between the change in CME on day 2 and change in CME on Day 5. Similar to our previously published data which consistent of half of the participants in the present data set [9] the ICC was in the good range (i.e. ICC between 0.6-0.8, ICC = 0.63). Furthermore, 77% of participants showed at least a 17% change in map volume, which in our previous study, was defined as a meaningful change in CME [9].

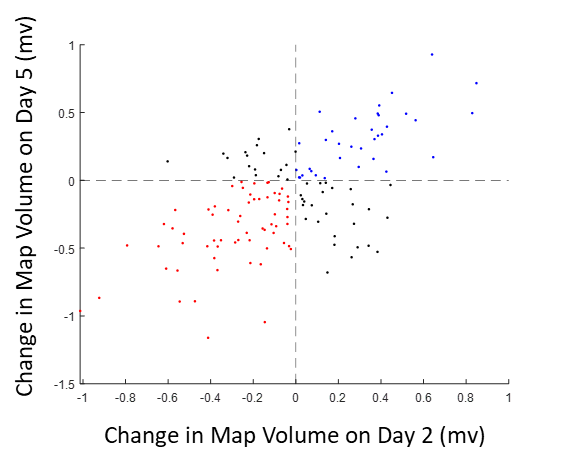

*eFigure 7.* Corticomotor map volume change scores on Day 2 plotted against change scores for Day 5. The blue plots show participants who demonstrated an increase in map volume on both Day 2 and 5, while the red plots show the opposite pattern. The orange plots show participants that demonstrate “unstable” changes i.e., a decrease on one day but an increase on the other.

**Pain Data Separated by Facilitator, Depressor, Fast PAF and Slow PAF**

eFigure 8 shows the pain scores (sum of chewing and yawning) across participants categorised either as depressor or facilitator (where map volume on Day 5 was respectively less or more than map volume on Day 0), and as having fast or slow PAF (respectively more or less than the median PAF value across participants).

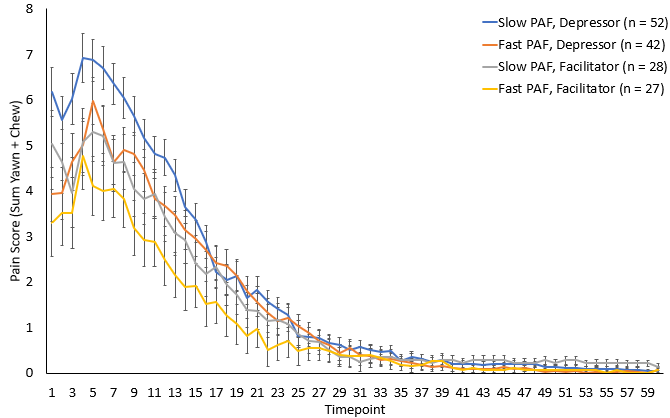

*eFigure 8*. Mean (± SEM) pain scores for participants categorised as having fast/slow PAF or a facilitator/depressor.

eFigure9 shows the difference in pain ratings between slow and fast PAF participants and between facilitators and depressors.

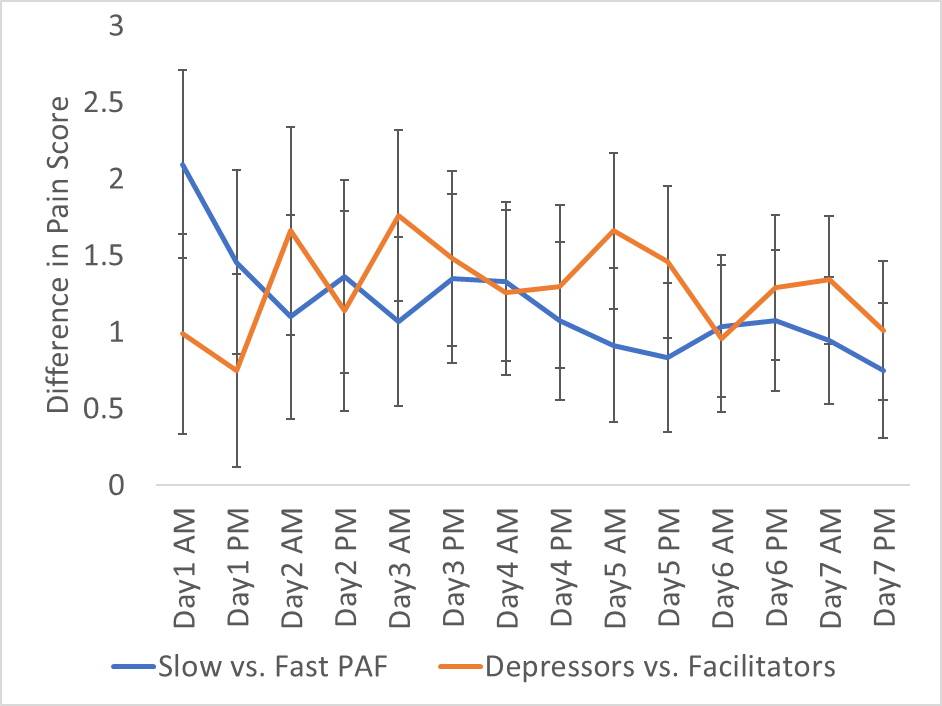

*eFigure 9.* Difference in pain ratings between slow and fast PAF participants and between facilitators and depressors. Error bars show the pooled standard error of pain scores for facilitators and depressors (orange line), and for slow and fast PAF participants (blue line).

**Additional Modelling Results**

eFigure 10 shows the training and test set ROCs when including PAF alone or CME alone in the predictive modelling.

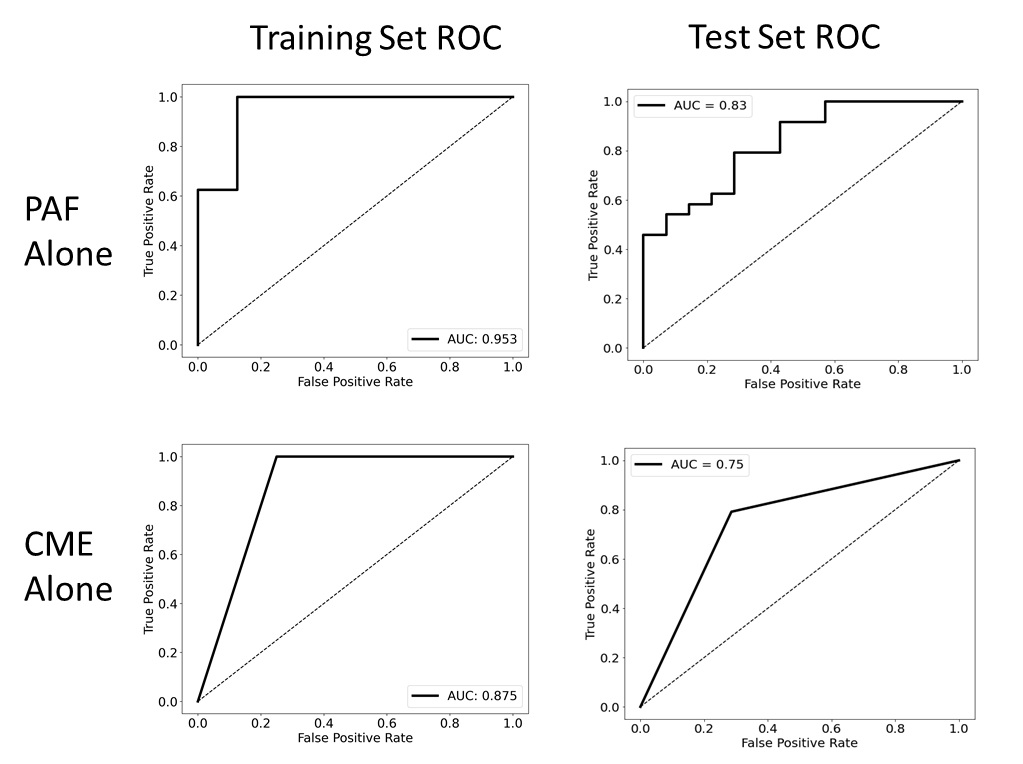

*eFigure 10.* Internal validation area under the curve (AUC) and Test Set AUCs when including peak alpha frequency (PAF) alone or corticomotor excitability (CME) alone in the predictive modelling.

eFigure 11 shows the training and test set ROCs when the pain diary data was not imputed through linear interpolation

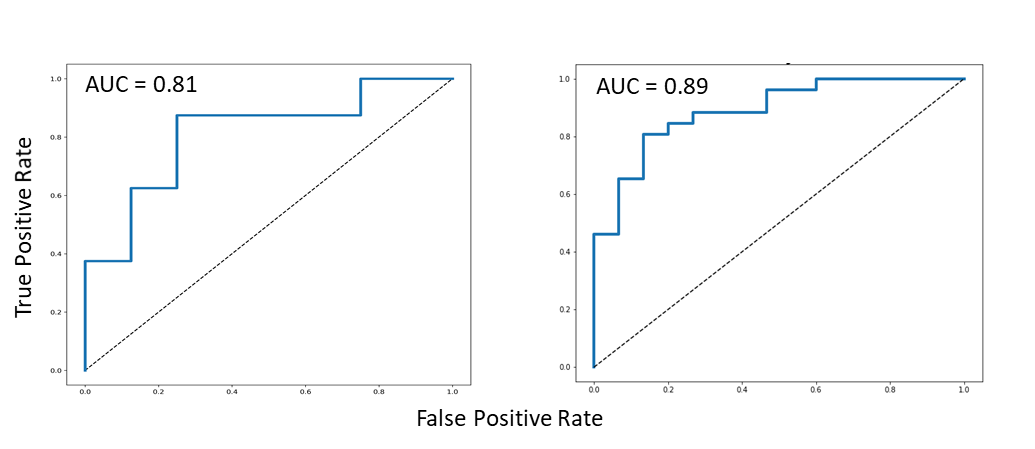

*eFigure 11.* Training validation area under the curve (AUC, left) and Test Set AUCs (right) when the pain diary data was not imputed through linear interpolation.

eFigure 12 shows the training and test set ROCs where latent growth modelling (LGM) pain labels were established using the whole 30 days of data (instead of only first 7 days).

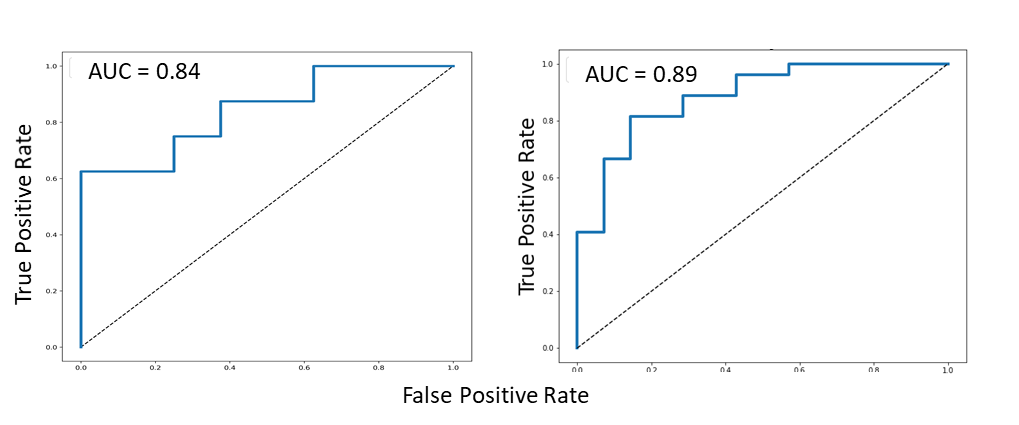

*eFigure 12.* Training validation area under the curve (AUC, left) and Test Set AUC (right) where latent growth modelling (LGM) pain labels were established using the whole 30 days of data (instead of only first 7 days).

eFigure 13 shows the training and test set ROCs when including CME was treated as a continuous variable i.e. CME = map volume for Day 5 – Day 0 divided by Day 0. The optimal probability threshold for detecting a high pain sensitive individual was 0.52. We found that when PAF was at an average value of 9.99Hz, a greater than 13% reduction in CME was the threshold for being labelled as high pain sensitive. When CME was at an average change score of +0.08%, a PAF value of less than 9.88 Hz was required to pass the threshold for being labelled as high pain sensitive.

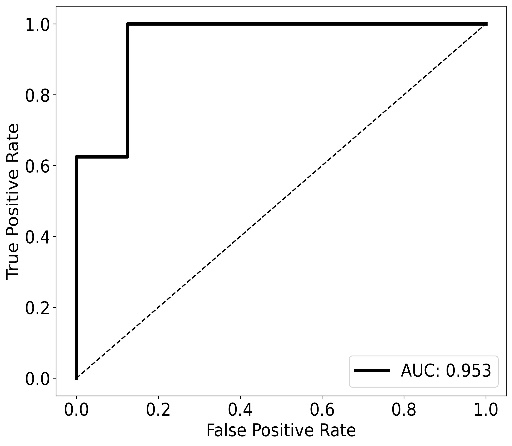

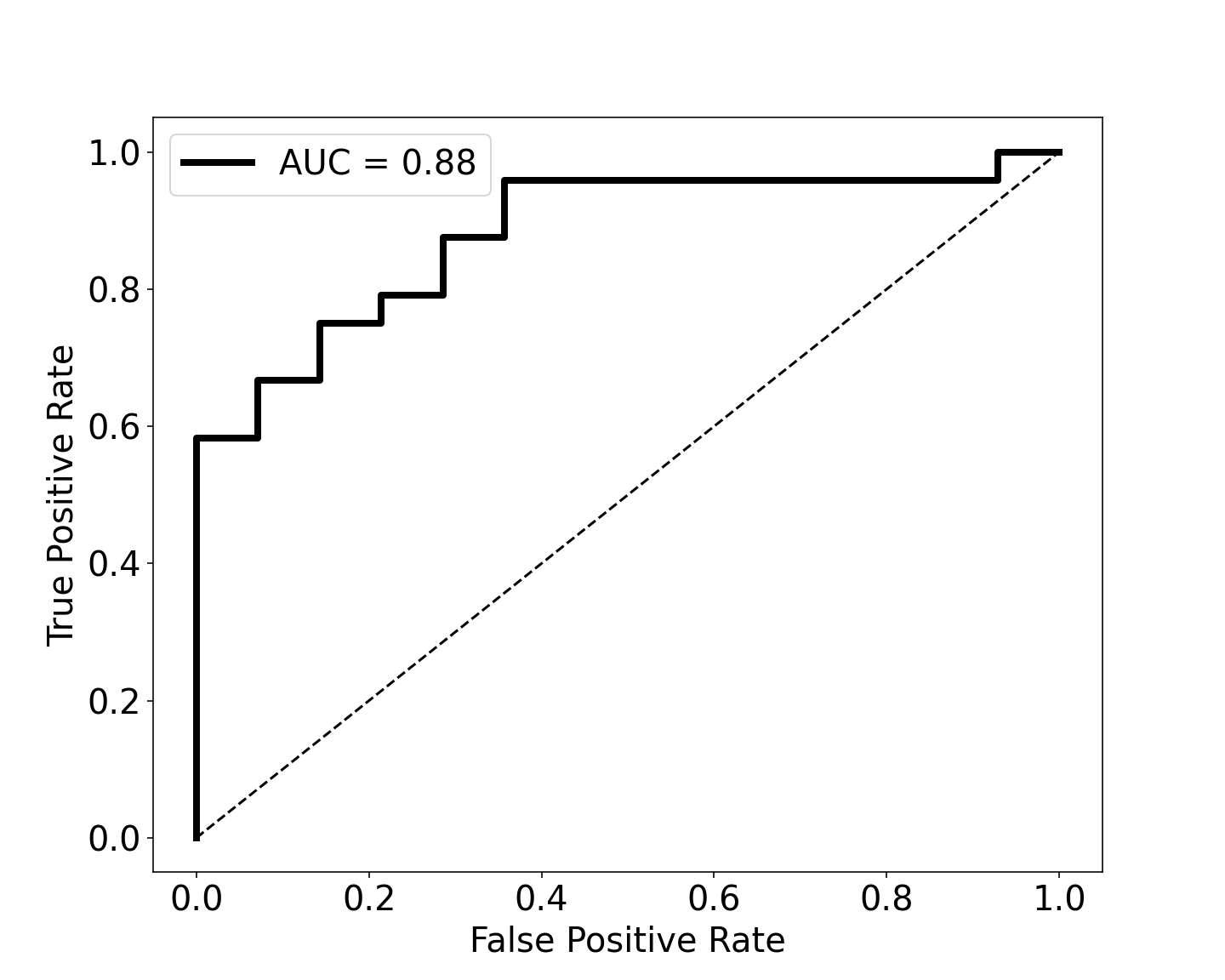

*eFigure 13.* Training validation area under the curve (AUC, left) and Test Set AUC (right) when CME was treated as a continuous variable (map volume for Day 5 – Day 0 divided by Day 0)
